# Strategies to enroll and retain low-income adolescent and young adult pregnant women in longitudinal studies: lessons learned from the AMOR project

**DOI:** 10.64898/2026.04.13.26350540

**Authors:** Saionara M. A. Câmara, Juliana Fernandes de Souza Barbosa, Sarah Hipp, Sabrina Gabrielle Gomes Fernandes Macêdo, Tetine Sentell, Diego G. Bassani, Marlos R. Domingues, Catherine M. Pirkle

## Abstract

**Background:** Prospective studies of pregnant adolescents are essencial to effectively address this global health priority. They help answer vital questions about their health, but such studies are uncommon due to the difficulty in retaining adolescents. This paper describes the successes and challenges of the research strategies used to ensure sufficient recruitment and retention of pregnant adolescents in a longitudinal study about adolescent childbearing in an under-resourced setting.

**Methods:** The Adolescence and Motherhood Research project was conducted in a rural region of Northeast Brazil in 2017-2019 and assessed 50 primigravids between 13-18 years (adolescents) and 50 primigravids between 23-28 years (young adults) during the first 16 weeks of pregnancy with two follow-ups (third trimester of pregnancy, and 4-6 weeks postpartum). Recruitment strategies involved engagement of health sector and community, as well as referrals from health care professionals and dissemination of the project in different locations. Retention strategies included maintaining contact with the participants between assessments and providing transportation for them to attend the follow-up procedures.

**Results:** Recruitment took 10 months to complete. A total of 78% of the participants were recruited from the primary health care units, mainly after referral from a health care provider. Retention reached 95% of the sample throughout the study (90%: adolescents; 98%: adults).

**Conclusion:** A combination of approaches is necessary to successfully recruit and retain youth in longitudinal studies and engaging local stakeholders may help to increase community-perceived legitimacy of the research. Working closely with front-line staff is essential when conducting research in rural low-income communities.

## Background

Important research questions focusing on adolescent fertility remain unanswered, despite evidence that adolescent pregnancy and childbirth is associated to long-term adverse health outcomes^1–3^. Complications from childbirth constitute the leading cause of death among girls ages 15-19 years in low- and middle-income countries (LMIC)^4,5^, and in Latin America (LA), half of the 1.2 million unplanned pregnancies occur among adolescents ^6^. The Latin America and Caribbean region has the second highest adolescent fertility rate of all world regions and research examining trends in 21 countries in the region showed that only about half have had statistically significant reductions in adolescent fertility across age groups, and some, have experienced increases^7^.

Despite the large number of adolescent pregnancies, research on adolescent fertility in LA is variable in quality and, in some countries, grossly insufficient^7^. Achievement of national programs to reduce adolescent fertility, as well as for multiple sustainable development goals that depend on lowering adolescent fertility^7^, requires a robust evidence base on the diverse drivers of adolescent fertility in the region. Progress in lowering adolescent fertility rates in LA is inequitable across and within countries; lower income, rural women, Indigenous and Afro-descendent women have disproportionately higher adolescent fertility rates compared to women that are better off, urban, and from other ethnicities ^7^. This highlights the importance of high quality research in disadvantages settings. One factor limiting research on adolescent fertility is the difficulty in recruiting and retaining pregnant adolescents in prospective research in LA, especially those from disadvantaged backgrounds^8,9^.

Prospective studies of adolescent pregnant women are viewed as difficult to implement, require extensive engagement strategies and the dedication of large research teams, making them expensive to conduct in comparison to other age groups ^8–11^. In addition, many institutional review boards consider under-age and pregnant women vulnerable populations^12^, requiring additional protections and administrative burden, while also adding liability issues^13^. In the United States, a vulnerable population indicates “vulnerability to coersion and undue influence” with regard to making an informed decision about participating in research^14^ (COMMON RULE). In Brazil, the National Health Council (N° 466/2012) defines the vulnerable population as “those whose capacity for self-determination is reduced or impaired, or who are in any way unable to resist, especially regarding free and informed consent”. This creates challenges in understanding and consequently leads to the exclusion of these individuals from research. Thus, excluding pregnant and underage women from research is a common strategy to simplify logistics and research bureaucracy ^15^. It should be acknolwedge that in 2019, the United States did remove the vulnerable population classification for pregnant women, but retains additional criteria for their inclusion in research studies that hampers research on this population ^13^. Underaged women remain a vulnerable population.

Pregnant women are also often excluded from research due to the (perceived) fetal risk associated with certain research procedures, the complicated physiology of pregnancy, and/or uncertainty about pregnant women’s willingness to participate ^10,16^. Yet, health during the periconceptional period and throughout pregnancy has life-long implications for the mother ^17–20^ and sets the developmental course of the fetus ^21^. Thus, it is a critically important life-stage to research. Exclusion from research studies only increases women and their infants’ vulnerability to poor health due to lack of evidence to support interventions ^22^. In addition to all of these issues, the stigma of adolescent pregnancy in many cultures may lead adolescents to hide the pregnancy, introducing another challenge ^23^. Prospective studies focusing on adolescent pregnancy are almost exclusively from urban areas ^24^. Subsequently, rural and remote contexts experience even larger gaps in quality evidence on adolescent pregnancy ^25^, despite adolescent pregnancy and childbirth being more common in rural regions in LA and elsewhere ^26,27^. In Brazil, the prevalence of childbirths among adolescent mothers is higher in rural than in urban areas ^28^ and these areas have distinct recruitment and retention challenges.

Given the limited resources describing strategies for engagement of pregnant women in research studies, especially those who are lower-income and/or rural, this paper describes the strategies and methods adopted during a successful pilot longitudinal study of adolescent pregnancy in a rural area of northeast Brazil, highlighting lessons learned, facilitators and remaining challenges.

## Methods

### AMOR Project

The Adolescence and Motherhood Research (AMOR) project established a research platform to examine pathways linking adolescent childbearing to long-term adverse health consequences, specifically risk factors for chronic disease ^17–19^. The AMOR project was a pilot prospective longitudinal study designed to test feasibility of a larger study on the topic ^29^. Given the challenges noted above, the feasibility of future longitudinal research on this topic in this region depends on the successful engagement of pregnant adolescents and young-adult mothers in longitudinal research.

The AMOR project was based in the Trairi region of Rio Grande do Norte state (Figure 1). Its main city Santa Cruz hosts a satellite campus of the Federal University of Rio Grande do Norte, the Faculty of Health Sciences of Trairi – FACISA. In addition to Santa Cruz, the project included four smaller rural towns within the region’s health service catchment area: Lajes Pintadas, São Bento do Trairi, Campo Redondo and Tangará. One in five births in the Trairi region are from mothers ages 15-19 years ^28^. The Northeast region of Brazil, and rural areas in general, present higher prevalence of non-communicable diseases (NCDs) such as diabetes and hypertension than other regions [^30^]. Thus, research in this region is needed to better understand health risks and consequences for pregnant women, especially adolescents.

**Figure 1.**
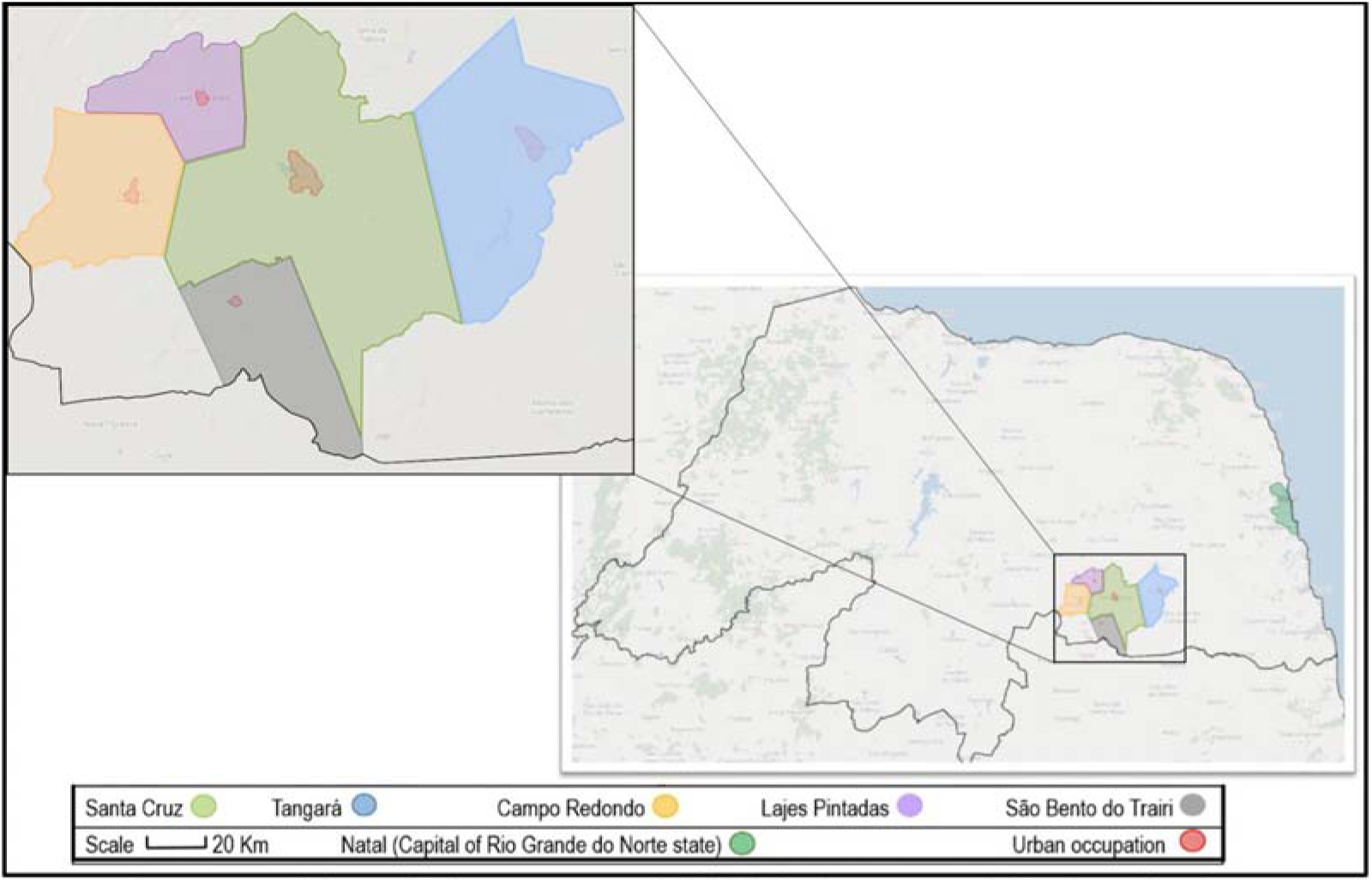
Map of Rio Grande do Norte state (Brazil) highlighting the AMOR study site (the Trairi region).

### Recruitment and retention strategies developed for the AMOR project

#### Study design

The study prospectively followed a sample of 50 adolescents (ages 13-18 years) and 50 young adults (ages 23-28 years). The age range for the adolescent group was based on previous research showing that childbirth-related risks decline dramatically after the age 18 ^31^. For the adult group, the age range was defined based on evidence suggesting this age group was at lowest childbirth-associated later-life disease risk ^32,33^.

Participants were recruited during the first 16 weeks of pregnancy and followed at two additional time points: during the third trimester (after 27 weeks of pregnancy), and between 4 and 6-weeks postpartum. Study procedures included face-to-face interviews and collection of urine and blood samples. Participants also completed validated clinical, anthropometric and physical performance measures. Baseline and follow-up assessments took 1.5 hours and 1 hour respectively (a notable amount of engagement from participants). Strategies of recruitment and retention are described below.

#### Recruitment sites

Primary health care is universally funded by the Brazilian government through the Family Health Strategy (FHS) and there is almost complete coverage of prenatal care, with most women seeking care well within the first 16 weeks of pregnancy ^34,35^.

Each FHS team is responsible for up 1,000 households and includes a physician, nurse, nurse assistant, and full-time community health agents that are geographically placed to maximize health assistance to the catchment area population. Community health agents are people who live in the community, having a more social than technical profile, representing the link between the professional team and the community and are knowledgeable of community needs ^36^. They visit each household within the catchment area, identify pregnancies and schedule the first prenatal appointment at the nearest primary health care unit (PHU). Alternatively, pregnant women can self-refer to the PHU and schedule a consultation. Because PHUs are the primary (self-referred) or secondary (community health agent referral) point of contact with the health care system, we selected them as the primary recruitment sites for the study.

### Health sector and community engagement strategies

#### Health sector engagement

To establish community trust and engage different partners in this project, the principal investigators and study coordinator visited all seven PHUs in Santa Cruz and Municipal Health Departments of the targeted towns before initiating recruitment activities. The visits included a presentation about AMOR, allowed investigators to respond to questions, and obtained formal support from PHU’s and health departments. The study team used the visit to identify points of contact at each PHU, as these health care providers would be required to maintain regular contact with the project coordinator throughout the study.

#### Community engagement

Given these rural communities’ lack of familiarity with health research activities, it was necessary to utilize a variety of approaches to create a context in which the study objectives could be understood and that established trust in the research team.

One of these activities involved introducing the study to the community and assuring them that it was authorized by the university, PHUs and other appropriate institutions. In Trairi, there is an extensive radio audience and history of collaboration between the radio and the university, with students and professors discussing health information and academic projects through broadcasts. The local principal investigator recorded two radio interviews, addressing the importance of adolescent pregnancy research and describing project objectives, which helped to establish project legitimacy, as the local population considers the radio a trustworthy source of information.

Informational posters and pamphlets were distributed in public spaces where potential participants would go including health centers, hospitals, clinics, schools, drug stores, fitness centers, grocery stores and beauty salons. These contained basic information about the project, such as the main objectives, inclusion criteria and contact information, so eligible participants could reach the research team. We used images and simple language (Supplemental files 1 and 2).

### Recruitment strategies

#### Referral from health professionals

Primary care providers were responsible for identifying eligible participants, based on age and first pregnancy. All front-line staff received a poster outlining inclusion criteria to be affixed to their desks to remind them about the project (Supplemental file 3). They were requested to inform potential participants about the project and ask if research staff could contact them to provide more details. Research staff weekly contacted each participating front-line provider, in person or by phone, to review the list of potentially eligible participants identified in the previous week or scheduled for an upcoming visit. Regular communication with health facility staff reminded them about the project and served to verify post-screening eligibility.

Participants were subsequently recruited directly by research staff. Brazilian ethics research regulations limits health care providers’ ability to engage in direct recruitment for research. Similarly, they do not allow researchers to provide participant incentives.

Due to the high volume of patients in certain PHUs in Santa Cruz, not all front-line staff were able to help with eligibility screening. Therefore, community health agents were tasked to assist with recruitment. Given the large number of community health agents in the region, weekly personal contact with each was impractical. Instead, study staff trained community health agents about the study protocol and recruitment details before initiating recruitment. In three of the five study towns, this was performed during large team meetings. Every week, during the recruitment period, we sent reminder text messages to alert them about eligible participants in their catchment areas.

For the large Santa Cruz PHUs, a research staff member monitored the waiting rooms during days on which prenatal appointments were offered. This visit served as an additional reminder about the project to providers and reduced their screening workload. Research staff explained the study and delivered pamphlets to potential participants in the waiting room. Pamphlets contained a description of the project and frequently asked questions, such as the criteria to participate, how often and when during pregnancy interviews would be performed, and basic information about study measurements and procedures (Supplemental files 1 and 2). At first contact, if the potential participant indicated her willingness to participate in the study, a private appointment was scheduled for the upcoming days. Because those under the age 18 needed ethical consent from their legal representatives according to Brazilian rules, the representatives were also provided with information about the study and invited to a private appointment. During this appointment, all participants and legal representatives of those under age 18 were informed of the study procedure details and follow-up assessments orally and with the consent and assent forms. The participants were also informed they could withdraw at any time.

Based on a co-author’s experiences implementing a birth cohort in Southern Brazil, which included pregnant women, we planned on including both male and female research staff as recruiters, but only female research staff as interviewers, as some participants are not comfortable being interviewed by men about pregnancy-related topics. Very early into the recruitment process we observed that potential participants were hesitant about the project, mainly when approached by male research staff, prompting us to adapt our strategy and use only female research staff for both screening and recruitment. Others researching reproductive health among rural youth have similarly reported less willingness to participate in a study, which in some cases resulted in refusal to participate or withdrawal from research after providing few responses ^37^. To address this concern, we believed that potential participants in AMOR would feel more comfortable talking to a peer. Therefore, our interviewers were young women ages 19-25. No participants withdrew from the study without completing at least the baseline interview.

### Retention strategies

Strategies to ensure recruited participants attended the two follow-up assessments began at the baseline interview. Participants were informed about upcoming evaluations and asked to provide all necessary information for future contact. Because changing telephone numbers and addresses are common in these communities, even within a relatively short time, we collected multiple types of contact information for the participants, including work telephone (mainly for the adults) or contact details of people closely related to the participant, such a parent or partner, as well as the participant’s home address, email, and social media contact.

An electronic scheduling software reminded study staff of each participant’s upcoming evaluations. Reminders were scheduled for 1-2 weeks before the follow-up window and, at that time, study staff sent a text message, using a freely available messaging service, to participants’ mobile phones informing them of an upcoming call from the study team to schedule the follow-up appointment. Finally, following a strategy described elsewhere [^38^], we sent personalized virtual cards on participant birthdays and after the child’s birth as a way to show our appreciation for study participation.

A study coordinator, who was not an interviewer, regularly verified participant satisfaction with our interviewers using a quality control questionnaire. The coordinator asked participants to give a grade from 5 to 10 for their interviewers’ professionalism and kindness during data collection, and to report if they had any discomfort or concerns related to the interviewers. The coordinators also explained that this was an independent assessment and that respondents of the quality control questionnaire would not be identified. All participants were contacted for this assessment after the baseline interview. We repeated this procedure with five randomly selected participants at each of the two follow-up interviews. Participant satisfaction was very high: all scores ranged from 8-10, and no participants reported discomfort or concern. During this contact, the study coordinator also asked a few other questions to check if responses matched the ones collected by the interviewers. The questions included information that would not change within the short time between data collection and quality control interview, such as date and place of birth, number of siblings, highest education level, and ever use of contraceptive. If at least one of the responses did not match the information previously collected, additional questions were asked. This served to identify concerns with the quality of the collected data and the need for additional training. No concerns were identified during all waves of data collection.

### Results and Discussion

#### Results, challenges and lessons learned

At the end of the recruitment process, a total of 207 pregnant women were contacted to obtain the desired sample size of 100 participants. The majority of exclusions were due to potential participants not meeting eligibility criteria: 57 women were not primigravids, 27 were identified after 16 weeks of pregnancy, 3 miscarried before baseline evaluation, 1 was prescribed bed rest due to placental abruption, 1 had a mental disorder, and 1 was living in a city outside the study area. Only 17 (14%) of the 117 eligible women declined participation. Table 1 presents the socioeconomic characteristics of the participants included in the study.

According to the Live Birth Information System of Brazil ^28^, from January to December of 2016, there were 217 adolescents (10-19 years old) who gave birth to a live child in the cities where this study occurred. Thus, we expected to identify approximately 18 pregnant adolescents in the region per month. Considering that adolescent pregnancy rates increase significantly at age 19, which was outside the inclusion age range, we hypothesized that it would require 6 to 8 months to recruit all participants, and that recruitment of the adolescent cohort would take longer than the young adult cohort. Surprisingly, recruitment of the adolescents was quicker than recruitment of adults, because adolescents were more likely to be primigravids. We needed 8 months to complete recruitment of the adolescent cohort, but 10 months to recruit the young-adult cohort. Almost half (47%) of the young-adult pregnant women we contacted were ineligible because they were not primigravids.

During the 8 months to recruit the adolescent sample, we contacted 81 individuals (only 8 of them were out of our age range). Considering the estimated rate of 18 new pregnant adolescents per month, we calculate that our recruitment strategies were able to identify, at a minimum, 56% of eligible pregnant adolescents in the region (81/144*100). We believe this underestimated of our actual recruitment of eligible adolescents, because we had no contact with many pregnant adolescents that we or the health care providers already knew were ineligible because they were out of our age range (e.g. 10 to 12 years and 19 years).

Although multiple strategies were used to advertise our study, 86% (N=43) of the adolescents and 64% (N=32) of the adults in our sample were recruited through referral by FHS teams during the weekly phone contact or during visits to PHUs. During visits to PHUs, we recruited another 3 (3%) participants who were not referred by staff, making the PHUs the main source of participants (N=78, 78%). Finally, 22 (22%) participants were recruited after word of mouth through a colleague or relative.

Our overall retention rate was 95% across the three waves of data collection; 90% for the adolescent group and 98% for the adult group. Only four participants (three adolescents and one adult) withdrew. We lost contact with one additional adolescent. Two adolescents had premature births (before 27 weeks) and were not assessed during third trimester of pregnancy. Although they missed the first follow-up assessment, both completed the postpartum evaluation. Figure 2 shows study retention.

The FHS team helped locate 3% (N=3) of the participants (all adolescents) for the follow-up assessments, since they could not be located using the information provided at baseline. These participants changed addresses during data collection and provided incorrect phone numbers.

**Figure. 2.**
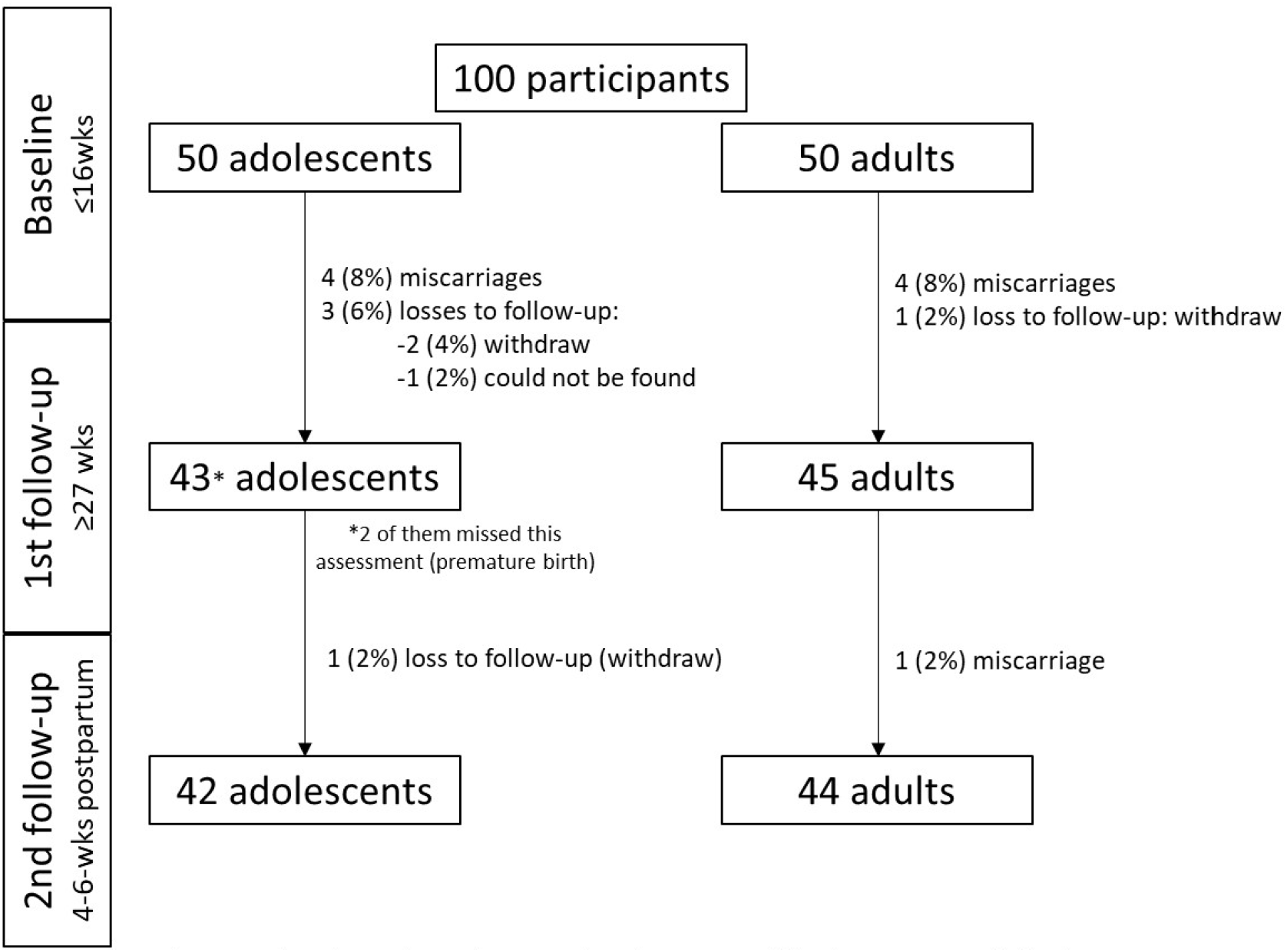
Flowchart of retention rates by cohort type within the AMOR study in the Trairi region, Brazil, 2017-2018.

Lack of public transportation and access in some rural communities were other important challenges during recruitment and follow-up. Thus, most assessments were performed at the PHU, normally located close to the participants’ residences. To guarantee the evaluation in a suitable setting and timeframe, we provided transport for 15 (15%) participants, mostly adolescents, from their homes to the PHU or university. Transport was offered when the participant reported that transport would be a barrier to participation and when we were unable to schedule interviews on the same day of their prenatal appointments. For the postpartum evaluation, we made arrangements to evaluate 54 participants (62% of the sample evaluated in this wave) at their residences, because of the additional difficulty to being out of home with a newborn and because they indicated feeling more comfortable staying at home.

## Conclusions

To successfully recruit and retain pregnant women from a rural region into our longitudinal study, a combination of approaches was necessary. The identification of potential participants by health care providers was the most important strategy (78% participants recruited this way) to reach our recruitment sample goals. Working closely with front-line staff and community health agents was essential, highlighting the importance of a good relationship between the research and health care teams. Because of providers’ workflows, flexible engagement strategies were required. We had very strong retention. Useful retention strategies included collecting multiple contact details and keeping in touch with participants during periods between evaluations. Some challenges required extra efforts and should be considered when planning longitudinal studies in similar contexts, such as lack of public transport and preference for female research staff. Despite the challenges faced and considering that incentives are not allowed in Brazil, our recruitment and retention showed a good response. The strategies and lessons learned from this project may prove useful for other research groups working with similar populations. They might also be useful for training researchers on how to plan and conduct a longitudinal study.

## Data Availability

All data produced in the present work are contained in the manuscript.

## Declarations

## Acknowledgements

The authors would also like to thank Ingrid Guerra Azevedo, PhD for her valuable support of this project.

## Ethics approval and Consent to participate

This study was approved by the following ethics committee: Comitê de Ética e Pesquisa da Faculdade de Ciências da Saúde do Trairi, Universidade Federal do Rio Grande no Norte (CEP-FACISA); Comissão Nacional de Ética em Pesquisa (CONEP); and Institutional Review Board of the University of Hawai□i.

## Consent for publication

Not applicable

## Availability of data and materials

Not applicable

## Author Contributions

SMAC and CMP conceptualized the project and led the implementation of the study. SMAC, JFSB, SH, SGGF and CMP drafted the manuscript. SMAC, TS, MRD, DGB and CMP developed the recruitment and retention procedures. All authors reviewed and approved the final version of the manuscript.

